# The Impact of Continuous Positive Airway Pressure Therapy on the Recurrence of Atrial Fibrillation in Patients with Obstructive Sleep Apnea after pulmonary vein isolation

**DOI:** 10.1101/2023.10.17.23297177

**Authors:** Jun Fan, Shao-Xi Sun, Li-li Cao, Shao-ling Luo, Shao-hua Wang, Wei-jie Li, Yi-chao Pan, Tian-yuan Wu, Jian Liu, Bing-Bo Yu

## Abstract

**Background:** Despite the recognized risk of atrial fibrillation (AF) recurrence after pulmonary vein isolation (PVI) in patients with obstructive sleep apnea (OSA), the effect of continuous positive airway pressure (CPAP) remains inconsistent across studies, necessitating further examination.

**Methods:** Utilizing databases encompassing Web of Science, Pubmed and OVID, we implemented a meta-analysis dedicated to investigating the role of OSA in post-PVI AF recurrence and the preventative properties of CPAP.

**Results:** Our meta-analysis of OSA patients undergoing PVI suggests an AF recurrence risk with a RR (risk ratio) of 1.67 (95% CI: 1.52-1.83). For patients with no atrial size difference, the risk is RR=2.13 (95% CI: 1.63-2.79), and with size difference, it’s RR=1.78 (95% CI: 1.46-2.17). Diagnoses from the Berlin questionnaire and polysomnography yielded RRs of 1.71 (95% CI: 1.37-2.14) and 1.75 (95% CI: 1.40-2.18), respectively. Non-CPAP usage increases AF recurrence risk by 67% and especially in cases of significant atrial size difference (RR=1.63, 95%CI:1.32-2.03). Conversely, when atrial size difference is absent, the impact of CPAP appears to be insignificant (RR=1.22, 95%CI: 0.98-3.02). Co-existence of paroxysmal and non-paroxysmal AF indicates a significant CPAP effect (RR=1.78, 95% CI: 1.50-2.09), contrary to one study on paroxysmal AF patients (RR=1.3, 95%CI: 0.71-1.50).

**Conclusion:** Our meta-analysis found a significant risk of AF recurrence in patients with OSA following PVI. However, OSA had an insignificant impact on AF recurrence in paroxysmal AF patients. Non-CPAP usage generally increased recurrence risk. Yet, in subgroups without prominent atrial size difference and paroxysmal AF, CPAP’s influence was not significant. Keywords: CPAP; Atrial Fibrillation; Obstructive Sleep Apnea; Pulmonary vein isolation

## Introduction

Pulmonary vein isolation (PVI) is widely recognized as a cornerstone treatment for atrial fibrillation (AF).^1^ Nonetheless, managing AF recurrence following PVI continues to present pressing and unresolved challenges in the clinical practice. The likelihood of such recurrence is known to vary broadly across different patient groups, ranging between 20%-50%.^2^ Several risk factors such as age, sex, AF type, structural heart disease, hypertension, and obstructive sleep apnea (OSA) have been recognized for their possible contribution to an elevated risk of AF recurrence following PVI.^3,4^ Therefore, effective management of these risk factors is instrumental in preventing AF recurrence. In patients with AF, OSA is of particular concern as a modifiable and treatable risk factor.^5^ However, the confluence of studies investigating the effectiveness of continuous positive airway pressure (CPAP) in preventing AF recurrence post-PVI has yielded disparate outcomes.^6,7^ Although earlier meta-analyses proposed a potential advantageous role for CPAP, more recent randomized controlled trials suggest a lack of significant difference in the effect of CPAP on AF recurrence.^8^ Consequently, the precise impact of CPAP on AF recurrence post-PVI warrants more detailed exploration.

Considering the current situation described previously, we embarked on this meta-analysis. The cardinal objectives of our study were twofold: (1) to scrutinize the implications of obstructive OSA on the recurrence of AF, whilst integrating the latest research insights into our investigation, and (2) to evaluate the efficacy of CPAP in mitigating the recurrence rate of AF amongst patients with OSA who underwent PVI.

## Methods

### Search strategy

The present study was registered on the Prospero website (Registration number: CRD42023452039, https://www.crd.york.ac.uk) in accordance with the guidelines specified by the Meta-analysis of observational studies in epidemiology (MOOSE) group. The databases searched for relevant studies included Pubmed, Web of Science, and Ovid. The search terms used in this study encompassed “atrial fibrillation”, “pulmonary vein isolation” and “obstructive sleep apnea”. Two investigators screened the retrieved results for eligibility.

### Study Selection

Only English-language literature was included in this study. “Conference proceedings,” “editorial viewpoints,” and “case reports” were excluded due to the lack of specific research details. The inclusion criteria for this study were as follows: 1) Participants had to be above 18 years of age; 2) prospective studies, retrospective studies, and case-control studies were eligible for inclusion; 3) For the aim of contrasting the influence of OSA on AF recurrence post-PVI intervention, comprehensive details relating to patients with OSA experiencing PVI therapy were necessitated; 4) When the study goal was to probe the repercussion of CPAP on post-PVI AF recurrence, treatment data focusing on CPAP in patients suffering from sleep apnea syndrome was investigated; and 5) a minimum follow-up period of 6 months was considered.

### Data Extraction and Quality Assessment

The data extracted from the literature in this study includes the title of the paper, publication date, country, type of research, number of patients, patients’ gender, age, BMI, comorbidities (such as hypertension, diabetes, etc.), types of AF (paroxysmal AF, persistent AF), methods of AF ablation, diagnostic methods for OSA (such as polysomnography, Berlin Questionnaire), and the post-ablation recurrence of AF in different populations (non-OSA population, OSA population without CPAP treatment, OSA population with CPAP treatment). The quality assessment of the included studies was performed using the “Newcastle-Ottawa” scale. Two investigators evaluated the selected literature. Any disagreements were resolved through discussion.

### Statistical analysis

Statistical analysis was performed using R software version 4.0.2 (R Foundation for Statistical Computing, Vienna, Austria) in this study. Continuous variables were expressed as mean ± standard deviation, while categorical variables were presented as numbers or percentages. Risk ratios (RR) accompanied by a 95% confidence interval were used to represent the risk assessment of AF recurrence. To quantitatively assess publication bias, Funnel Plot and Egger’s test were employed. A funnel plot was constructed for visual assessment of publication bias, while Egger’s test determined the presence of publication bias based on a significant p-value (<0.05). Heterogeneity testing was carried out using both the Q test and I^2^ test for each study. The Chi-square test was used to assess whether there was a significant statistical difference in I^2^, with a p-value less than 0.1 indicating such a difference. Where the I^2^ value was between 0-40%, no publication bias was inferred, guiding the implementation of a fixed-effect model. On the other hand, an I^2^ value surpassing 40% indicated the existence of publication bias, calling for the utilization of a random-effects model. The Mantel-Haenszel model was utilized to calculate the RR value, which incorporated the results from each study.

## Results

We initially retrieved a total of 970 articles based on our search criteria, as shown in Figure 1. After detecting and excluding duplicate articles (n=604), we conducted a rigorous review of the titles and abstracts of the remaining articles, leading to the further exclusion of 157 articles. The remaining articles were then subjected to a comprehensive full-text review. Among these, 3 articles were omitted based on specific exclusion criteria, as detailed in Figure 1. At the conclusion of this meticulous screening process, 13 articles were considered eligible and included in our study.

**Figure 1.**
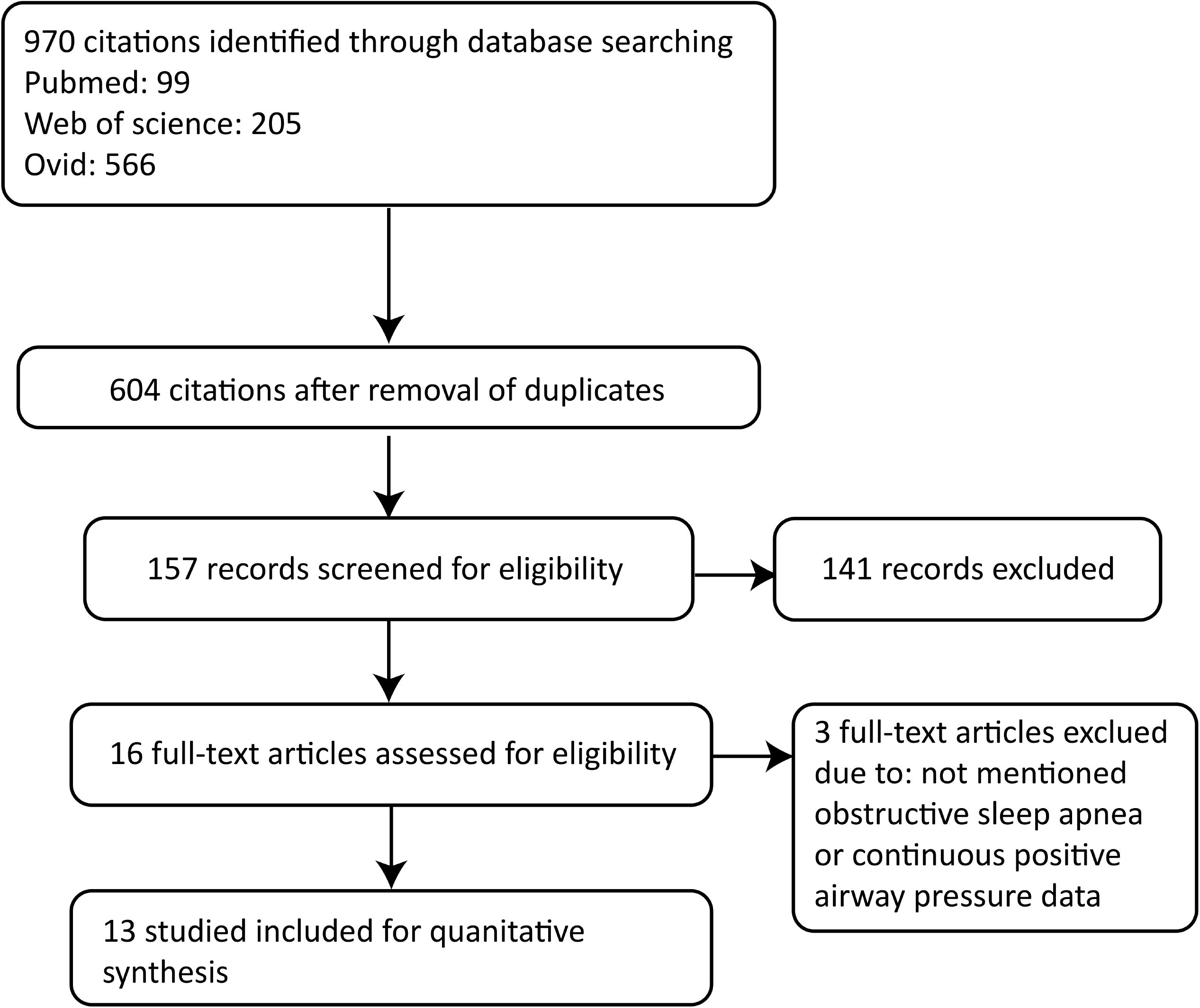
Flowchart depicting the study selection process in our meta-analysis.

Our selection of 13 articles included data from 4919 patients who underwent PVI for AF. Among these, 3910 individuals were not diagnosed with OSA, while 1004 individuals were identified as OSA patients. Of the latter group, 603 patients underwent CPAP treatment, while the remaining 628 patients did not receive CPAP therapy. The duration of follow-up across these studies varied, ranging from a minimum of 7 months to a maximum of 32 months. The characteristics and quality assessment of the included studies was showed in Table 1, while Table 2 presents the basic demographic details of the patients.

**Table 1.** Summary of the characteristics of included studies.

| Investigator | Year Published | Geographical Aera | Study Method | Enrollment of Patients (n) | AF Type at Enrollment | AF treatment | OSA diagnostic method | OSA treatment | Interval of follow-up (months) | Quality score |
| --- | --- | --- | --- | --- | --- | --- | --- | --- | --- | --- |
| Zhou<sup>17</sup> | 2022 | China | Prospective | 182 | P = 18<br>Ps = 144 | PVI | PSG | CPAP | 12 | 8 |
| Tang<sup>18</sup> | 2008 | China | Prospective | 178 | P = 178 | PVI | BQ | - | 12 | 9 |
| Szymanski<sup>19</sup> | 2015 | Poland | Prospective | 290 | P = 176<br>Non-P = 114 | PVI | PSG | - | 12 | 8 |
| Patel<sup>7</sup> | 2010 | America | Prospective | 3000 | P = 1603<br>Non-P = 1397 | PVI plus left atrial linear ablation | PSG | CPAP | 32 | 9 |
| Neilan<sup>20</sup> | 2015 | Netherland | Prospective | 720 | P = 249<br>Ps = 471 | PVI plus CAFE ablation | PSG | CPAP | 42 | 9 |
| Jongnarangsin<sup>21</sup> | 2008 | America | Prospective | 324 | P = 243<br>Non-P = 81 | PVI plus CAFE ablation | PSG | CPAP | 7 | 7 |
| Fein<sup>22</sup> | 2012 | America | Prospective | 114 | P = 53<br>Ps = 61 | PVI | PSG | CPAP | 12 | 9 |
| Chilukuri<sup>23</sup> | 2009 | America | Prospective | 210 | P = 119<br>Non-P = 91 | PVI | BQ | - | 25 | 9 |
| Chilukuri<sup>24</sup> | 2010 | America | Prospective | 109 | P = 84<br>Non-P = 25 | PVI | BQ | - | 11 | 9 |
| Hunt<sup>6</sup> | 2022 | Norway | RCT | 83 | P = 83 | PVI | PSG | CPAP | 12 | 9 |
| Naruse<sup>4</sup> | 2012 | Japan | Prospective | 153 | P = 82<br>Non-P = 71 | PVI plus left atrial linear ablation or superior vena cava ablation | PSG | CPAP | 18 | 9 |
| Hojo<sup>25</sup> | 2018 | Japan | Prospective | 100 | P = 89<br>Non-P = 11 | PVI | PSG | CPAP | 24 | 8 |
| Matiello<sup>26</sup> | 2010 | Spain | Prospective | 174 | P = 98 | PVI plus | BQ | CPAP | 17 | 8 |
|  |  |  |  |  | Non-P = 76 | left atrial<br>linear<br>ablation |  |  |  |  |

BQ, Berlin questionnaire; CAFE, complex fractionated atrial electrogram; PSG, polysomnography; PVI, pulmonary vein isolation; AF, atrial fibrillation; OSA, obstructive sleep apnea; (-), data not available; RCT, randomized controlled trial; P, paroxysmal; Ps, persistent.

**Table 2.** Patient characteristics of included studies.

| Primary investigators | Year Published | OSA |  |  |  |  | Non-OSA |  |  |  |
| --- | --- | --- | --- | --- | --- | --- | --- | --- | --- | --- |
|  |  | Patients | Age(years) | CPAP Patients | HT | DM | Patients | Age(years) | HT | DM |
| Zhou<sup>17</sup> | 2022 | 182 | - | 62 | 119 | 76 | 60 | 61 | 42 | 40 |
| Tang<sup>18</sup> | 2008 | 104 | 58 | - | 70 | 17 | 19 | 56 | 17 | 5 |
| Szymanski<sup>19</sup> | 2015 | 115 | 60 | - | 91 | 12 | 136 | 56 | 92 | 9 |
| Patel<sup>7</sup> | 2010 | 640 | 51 | 315 | 224 | 116 | 2360 | 57 | 1062 | 283 |
| Neilan<sup>20</sup> | 2015 | 142 | 57 | 71 | 90 | 33 | 578 | 56 | 275 | 73 |
| Jongnarangsin<sup>21</sup> | 2008 | 32 | 59 | 18 | 23 | - | 292 | 57 | 127 | - |
| Fein<sup>22</sup> | 2012 | 62 | - | 32 | 42 | 12 | 30 | 59 | 21 | 6 |
| Chilukuri<sup>23</sup> | 2009 | 92 | 58 | - | 63 | 9 | 118 | 58 | 43 | 10 |
| Chilukuri<sup>24</sup> | 2010 | 48 | - | - | - | - | 61 | - | - | - |
| Hunt<sup>6</sup> | 2022 | 83 | 62 | 37 | 32 | 4 | 21 | - | - | - |
| Hojo<sup>25</sup> | 2018 | 34 | - | 11 | 24 | 10 | 66 | 60 | 25 | 4 |
| Naruse<sup>4</sup> | 2012 | 116 | - | 82 | - | - | 37 | - | - | - |
| Matiello<sup>26</sup> | 2010 | 42 | - | - | 28 | - | 132 | 51 | 45 | - |

HT, hypertension; DM, diabetes mellitus; (-), data not available; OSA, obstructive sleep apnea; CPAP, continuous positive airway pressure

In our meta-analysis, we incorporated a total of 13 studies to explore the influence of OSA on the recurrence of AF after catheter ablation. Of these, 11 studies identified OSA as a risk factor for AF recurrence post radiofrequency (RF) ablation, while 3 studies suggested OSA had no significant influence on AF recurrence. The observed heterogeneity among the studies, represented by an I^2^ value of 56%, necessitated employing a random-effects model. On pooling the results from all studies, the RR for AF recurrence in the presence of OSA was found to be 1.67 [95% confidence interval (CI): 1.52-1.83] (Figure 2A). Sensitivity analyses further corroborated OSA as a risk factor for AF recurrence, unaffected by the removal of any particular study (Figure 2B). The publication bias for our selected studies is depicted in Supplementary Figure 1.

**Figure 2.**
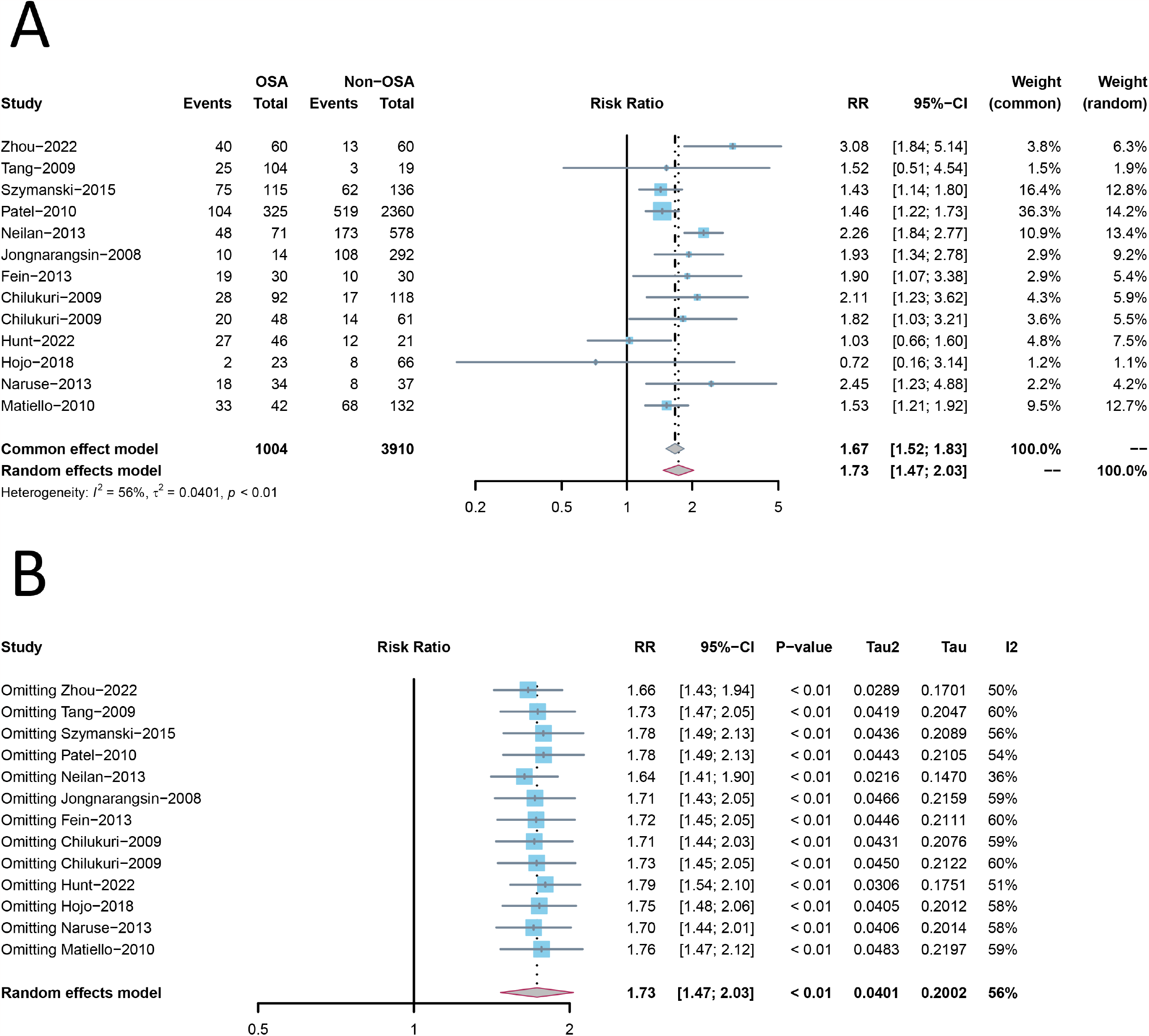
A. Forest plot comparing the recurrence rate of atrial fibrillation following radiofrequency ablation in patients suffering from obstructive sleep apnea (OSA) and those not suffering from OSA. B. Sensitivity analysis of recurrence rates of atrial fibrillation post-radiofrequency ablation in patients with OSA and those without OSA. OSA, obstructive sleep apnea.

We conducted a subgroup analysis based on whether there was a difference in the size of the atria. For the group without atrial size differences, the recurrence risk (RR) of AF in OSA patients stood at 2.13 (95% CI: 1.63-2.79) as illustrated in Figure 3. On the other hand, for the group presenting atrial size differences, this risk was evidenced at 1.78 (95% CI: 1.46-2.17). Sensitivity analysis verified that OSA remained a risk factor for AF recurrence even after the removal of any single study (Supplementary Figure 2). The diagnostic methods for OSA included the Berlin questionnaire and the polysomnography, with RRs of 1.71 (95% CI: 1.37-2.14) and 1.75 (95% CI: 1.40-2.18) respectively. There was insignificant effect of OSA on AF recurrence in patients with paroxysmal AF (RR=1.15, 95% CI: 0.74-1.77), but in the group with co-existing paroxysmal and persistent AF, the RR was 1.7 (95% CI: 1.54-2.11). Sensitivity analysis suggested that removing any single study did not affect the impact of OSA on post-ablation AF recurrence (Supplementary Figure 6).

**Figure 3.**
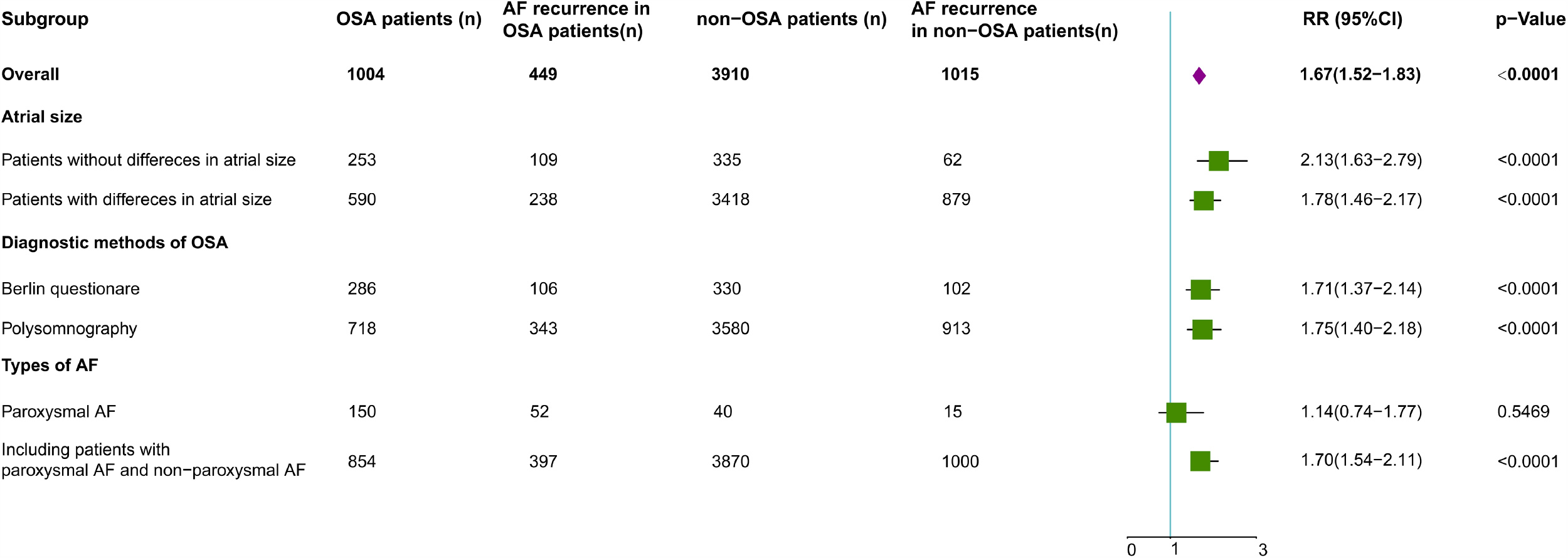
Subgroup analysis of recurrence rates of atrial fibrillation post-radiofrequency ablation in patients with OSA and those without OSA. AF, atrial fibrillation; OSA, obstructive sleep apnea.

We evaluated the impact of CPAP on recurrence in individuals with OSA. Our findings suggest that non-CPAP usage elevates the risk of AF recurrence in OSA patients by 67% (Figure 4). Sensitivity analyses reinforced these observations and demonstrated that the results consistently held while disregarding any arbitrary study.

**Figure 4.**
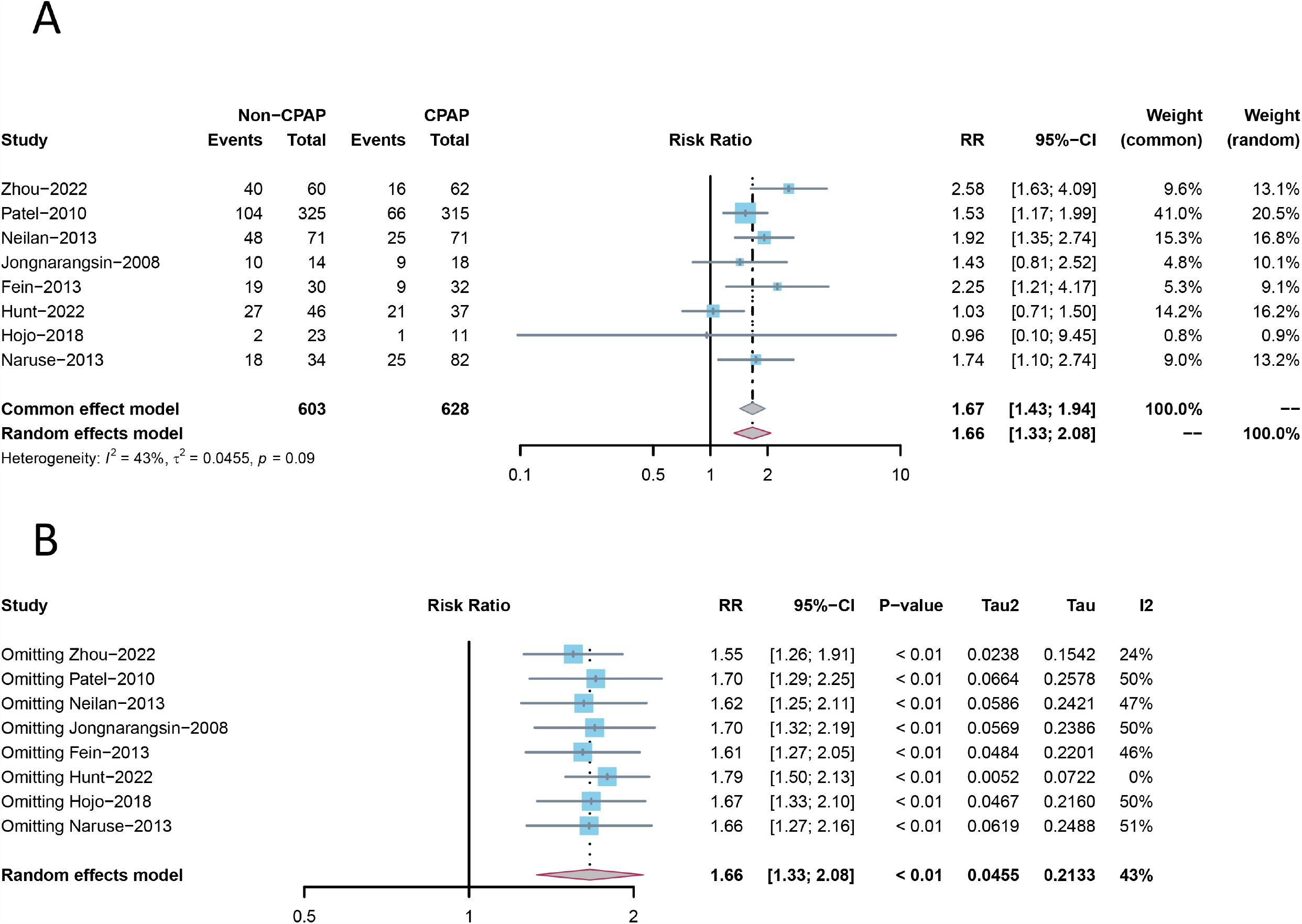
A. Forest plot exploring the recurrence of AF after radiofrequency ablation in OSA patients with and without CPAP usage. B. Sensitivity analysis graph evaluating the occurrence of AF recurrence after radiofrequency ablation in OSA patients, comparing those using CPAP and those not. OSA, obstructive sleep apnea.

Our subsequent subgroup analyses, performed based on size of atrium, indicated no significant impact of CPAP on AF recurrence post-ablation (RR=1.22, 95%CI: 0.98-3.02) when no prominent difference in the atrial size was observed (Figure 5A). When discounting any other arbitrary studies, sensitivity analyses have indicated that non-CPAP usage does not significantly affect the recurrence of AF, with the exception of the research conducted by Hunt et al^6^ (Supplementary Figure 9A). When significant difference in atrial size was observed among the patients involved in the study, not using CPAP was found to be a risk factor for recurrent AF (RR=1.63, 95%CI:1.32-2.03) (Figure 5B). Additional subgroup analyses were undertaken on studies that exclusively involved patients with paroxysmal AF and those which incorporated both patients with paroxysmal AF and non-paroxysmal AF. When both paroxysmal and non-paroxysmal AF were present in the participants, the RR value for the impact of non-CPAP usage on AF recurrence escalated to 1.78, 95%CI: 1.50-2.09 (Figure 5C). Nevertheless, A single study found no substantial impact of CPAP on post-ablation AF recurrence in patients that showed paroxysmal AF, with a RR of 1.3, 95%CI: 0.71-1.50 (Figure 5D). Consistent results were yielded by sensitivity analyses.

**Figure 5.**
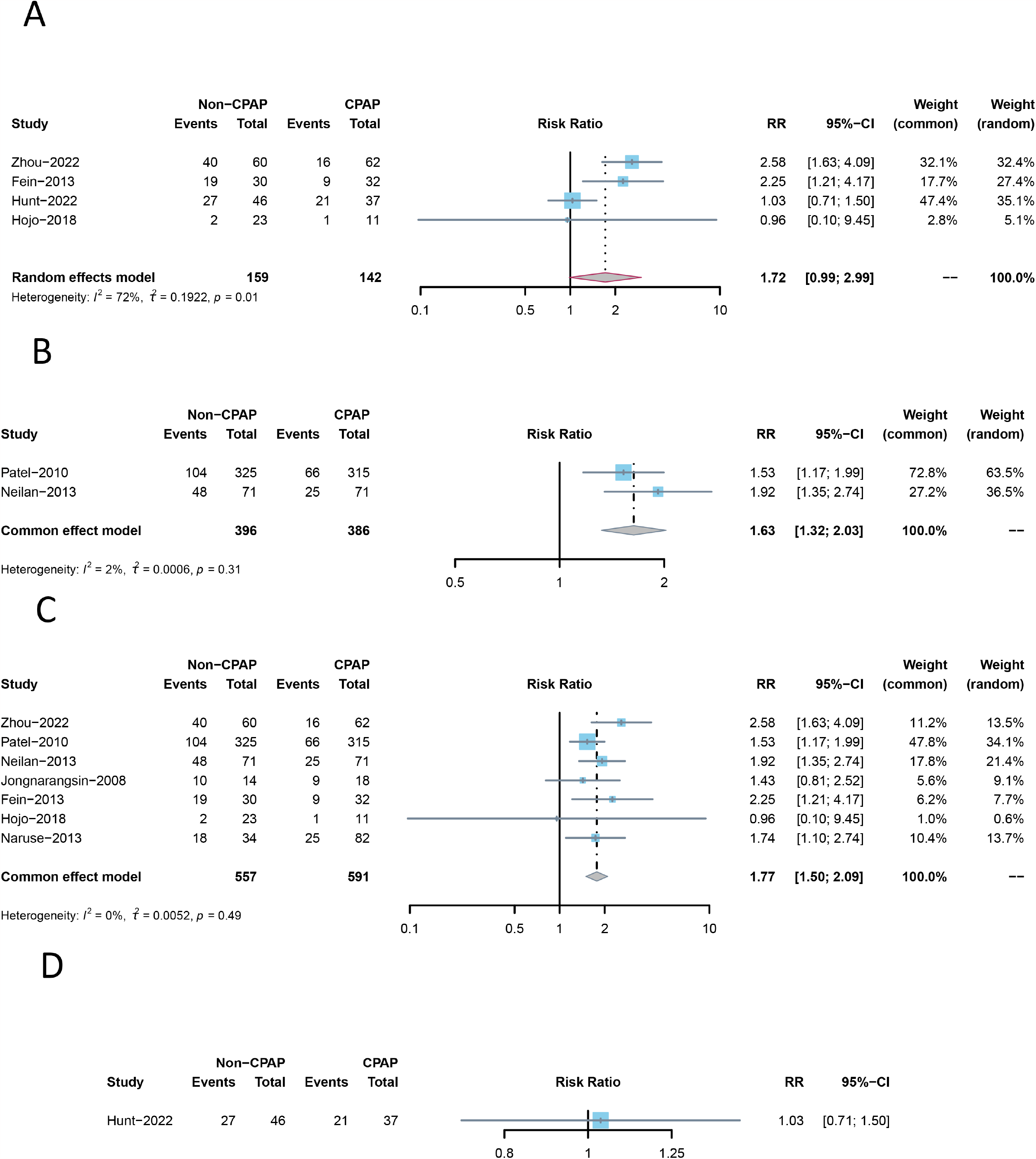
Subgroup analysis on AF recurrence following radiofrequency ablation in OSA patients, categorized by CPAP usage. A. No statistical atrial size difference in CPAP users and non-users among the included studies. B. A significant atrial size difference noted. C. Both paroxysmal and non-paroxysmal AF patients included in either group. D. Only paroxysmal AF patients included. OSA, obstructive sleep apnea.

## Discussion

In our comprehensive meta-analysis, we strived to elucidate the influence of OSA on the recurrence of AF post-catheter ablation. Our evaluation identified OSA as a contributory risk factor towards the recurrence of AF, with this conjecture being further reinforced by both sensitivity and subgroup analyses. Non-use of CPAP therapy in OSA patients was also linked with a higher AF recurrence risk. However, in instances where there is no substantial variation in the atrial size amongst patients, the use of CPAP negligibly affects the rate of AF recurrence post-ablation, as endorsed by our consistent sensitivity analyses. For patients with paroxysmal AF or concurrently paroxysmal and persistent AF, CPAP showed varied effects on AF recurrence post-ablation.

The association between OSA and the onset of AF has been confirmed by several studies^9^, a finding that our research further supports. OSA can promote the occurrence of AF through a variety of mechanisms, one of the most important of which is alterations in cardiac structure^10^. OSA-induced obstruction of the upper respiratory tract leads to significant variations in thoracic pressure, which substantially increase the difference in pressure inside and outside the heart chambers, eventually leading to atrial dilation^11^. Enlarged heart chambers can cause changes in the overall structure and fibrous makeup of the heart, thereby promoting the occurrence and progression of AF^12^. Compared to the control group, patients with OSA typically exhibit larger atrial sizes and more severe left ventricular diastolic dysfunction.^13^

An enlarged atrial size has been identified as a risk factor for the onset of AF, a view supported by numerous studies^14^. Our research also underscores this conclusion, as we found that the frequency of AF recurrence after PVI significantly increased when the atrial size parameters in the OSA group were substantially larger than those in the non-OSA group. However, an important point underscored by our study is that the probability of AF recurrence in the OSA group is still higher than that in the non-OSA group even when there is no significant difference in atrial size parameters between the two. This suggests that in addition to the mechanism of altered cardiac structure promoting AF, OSA also involves other mechanisms that promote the occurrence and progression of AF, such as endothelial system dysfunction, inflammatory response, and oxidative stress.^15^

While polysomnography remains the primary diagnostic method for OSA, the user-friendly Berlin questionnaire has emerged as a significant adjunct in clinical practice. In an earlier meta-analysis, Chee Yuan Ng et al. discovered the Berlin questionnaire effectively identifies the risk of recurrence of AF (AF) in patients with OSA following PVI.^16^ This finding was subsequently corroborated by my research. These discoveries further attest to the practicality and efficacy of the Berlin questionnaire in anticipating the risk of postoperative AF recurrence in OSA patients.

Our research findings suggest that there is no significant difference in the influence of OSA on the recurrence of AF among patients with paroxysmal AF compared to those without OSA. However, when the study population includes both paroxysmal and persistent AF patients, OSA prominently emerges as a notable risk factor for the recurrence of AF. Existing literature does not provide a detailed explanation for this phenomenon. We hypothesize that this may be attributable to minimal differences in atrial structural alterations between OSA and non-OSA patients among those with paroxysmal AF, resulting in no significant divergence in their risk of recurrence.

Consistent with previous meta-analysis results, our study further confirms that CPAP can effectively reduce the risk of AF recurrence after PVI.^8^ However, when the atrial size of the patients participating in the study showed no significant differences, we found that the effect of CPAP in preventing recurrence of AF was not significant. Similarly, there was not a significant difference found in the impact of CPAP among patients with paroxysmal AF. Regarding this phenomenon, a definitive explanation was not identified from our review of related literature. We speculate that this observation could be related to the reasons we have previously mentioned, i.e., the atrial structural changes are likely minor among OSA patients with paroxysmal AF and those without significant changes in atrial size, which might be the primary reason for the lack of significant differences in recurrence rates. Therefore, the understanding of the influence of CPAP on AF recurrence requires reevaluation, and more clinical trials are needed to further substantiate this point of view.

### Limitations

In our meta-analysis, all data were not adjusted for cardiovascular related risk factors, including gender, age, baseline diseases (such as hypertension, diabetes, etc.), and baseline medication status. Furthermore, our study did not differentiate the severity of OSA. Additionally, there was substantial heterogeneity among the studies in some of the comparisons, which might have influenced the results. Lastly, most of the results selected for our study were from retrospective studies, potentially leading to bias between the experimental and control groups, thus potentially affecting the study outcomes.

## Conclusions

The conclusion of our meta-analysis highlighted OSA as a risk factor for the recurrence of atrial AF following catheter ablation. Our analysis also revealed that OSA patients who do not use CPAP therapy are at a higher risk of AF recurrence. However, for patients with no significant variation in atrial size, the use of CPAP had no effect on the rate of AF recurrence post-ablation. The impact of CPAP on AF recurrence varied among patients with paroxysmal AF or co-existing paroxysmal and persistent AF.

## Data Availability

If authorized by the author, the data may be available upon request.

## Compliance with Ethical Standards

Conflict of Interest The authors declare that they have no conflict of interest.

## Human Subjects/Informed Consent Statement

This study has been approved by the Ethics Committee of the Second Affiliated Hospital of South China University of Technology. It was conducted in accordance with the Helsinki Declaration of 1975 (revised in year 2000).

## Figure legends

Supplementary Figure 1. Funnel plot comparing the recurrence rate of atrial fibrillation after radiofrequency ablation in patients with obstructive sleep apnea (OSA) and those without OSA.

Supplementary Figure 2. This illustration shows a sensitivity analysis comparing the recidivism of atrial fibrillation following radiofrequency ablation procedures between patients with Obstructive Sleep Apnea (OSA) and those without. Two situations are analyzed: (A) where the enrolled patients exhibit no significant disparity in their atrial sizes and (B) where a substantial difference in atrial sizes among patients is observed.

Supplementary Figure 3. Funnel plots of studies which illustrate recurrence rates of atrial fibrillation post-radiofrequency ablation in patients with and without Obstructive Sleep Apnea (OSA), when there was no significant difference noted in the atrial sizes (A) and significant variation in atrial sizes (B).

Supplementary Figure 4. This illustration depicts a sensitivity analysis which compares the recurrence rate of atrial fibrillation in patient’s post-radiofrequency ablation, between those with Obstructive Sleep Apnea (OSA) and those without. Two diagnostic methods are analyzed: (A) using the Berlin questionnaire and (B) using the polysomnography.

Supplementary Figure 5. These funnel plots of studies illustrated the recurrence rates of atrial fibrillation post-radiofrequency ablation in patients with and without Obstructive Sleep Apnea (OSA), diagnosed via (A) the Berlin questionnaire and (B) polysomnography.

Supplementary Figure 6. This illustration demonstrates a sensitivity analysis comparing the recurrence rates of atrial fibrillation in patients post-radiofrequency ablation, distinguishing between those with Obstructive Sleep Apnea (OSA) and those without. Included research involves: (A) studies which involve patients with paroxysmal atrial fibrillation and (B) studies that have patients with both paroxysmal and non-paroxysmal atrial fibrillation.

Supplementary Figure 7. Funnel plots of studies presenting the recurrence rates of atrial fibrillation post-radiofrequency ablation in patients with and without Obstructive Sleep Apnea (OSA). The panels include: (A) studies which patients have paroxysmal atrial fibrillation and (B) studies involving patients with both paroxysmal and non-paroxysmal atrial fibrillation.

Supplementary Figure 8. Funnel plot of studies included that explore the occurrence of AF recurrence after radiofrequency ablation in OSA patients using CPAP and those not using CPAP.

Supplementary Figure 9. Subgroup analysis of studies included that explore the occurrence of AF recurrence after radiofrequency ablation in OSA patients using and not using CPAP. Two situations are analyzed: (A) where the enrolled patients show no significant disparity in their atrial sizes, and (B) where a substantial difference in atrial sizes among patients is observed.

Supplementary Figure 10. This funnel plot illustrates the array of studies undertaken to examine the incidence of atrial fibrillation (AF) recurrence following radiofrequency ablation in patients with Obstructive Sleep Apnea (OSA), categorized by the use and non-use of Continuous Positive Airway Pressure (CPAP). This visual representation demonstrates two distinct scenarios; (A) embodies a population of patients were negligible disparity in atrial dimensions is detected, whereas (B) depicts a situation that encompasses a pronounced discrepancy in atrial size among the study participants.

Supplementary Figure 11. A. This figure illustrates a sensitivity analysis focusing on the incidence of AF recurrence post-radiofrequency ablation in OSA patients, both using and not using CPAP. This analysis includes subgroup studies of patients with paroxysmal and non-paroxysmal AF. B. A funnel plot representation of the studies included in the research.

